# The impact of public health interventions on the future prevalence of ESBL-producing *Klebsiella pneumoniae*: a population based mathematical modelling study

**DOI:** 10.1101/19012765

**Authors:** Luisa Salazar-Vizcaya, Andrew Atkinson, Andreas Kronenberg, Catherine Plüss-Suard, Roger Kouyos, Viacheslav Kachalov, Nicolas Troillet, Jonas Marschall, Rami Sommerstein

## Abstract

**Background:** Extended-spectrum betalactamase (ESBL-) producing *K. pneumoniae* is one of the most common causes of infections with antimicrobial resistant bacteria worldwide. The spread of colonization of humans with this pathogen is on the rise. The future prevalence of colonization with ESBL-producing *K. pneumoniae*, and the potential of public health interventions to lower it, remain uncertain.

**Methods:** Based on detailed data on antimicrobial consumption and susceptibility systematically recorded for over 13 years in a Swiss region, we developed a mathematical model to *i)* reconstruct the observed course of colonization with ESBL-producing *K. pneumoniae;* and *ii)* to assess the potential impact of public health interventions on future trends in colonization.

**Results:** Simulated prevalence of colonization with ESBL-producing *K. pneumoniae* stabilized in the near future when rates of antimicrobial consumption and in-hospital transmission remained stable in the main analyses (simulated prevalence in 2025 was 5.3% (5.0%-9.1%) in hospitals and 2.7% (2.1%-4.6%) in the community *versus* 5.6% (5.1%-9.5%) and 2.8% (2.2%-5.0%) in 2019). The largest changes in future prevalence were observed in simulations that assumed changes in overall antimicrobial consumption. When overall antimicrobial consumption was set to decrease by 50%, prevalence in 2025 declined by 89% in hospitals and by 84% in the community. A 50% decline in transmission rate within hospitals led to a reduction in prevalence of 43% in hospitals and of 13% in the community by 2025. Prevalence changed much less (≤9%) across scenarios with reduced carbapenem consumption. Assuming higher rates for the contribution from external sources of colonization, led to decreasing estimations of future prevalence in hospitals. While high uncertainty remains on the magnitude of these contribution, the best model fit suggested that as much as 46% (95% CI: 12%-96 %) of observed colonizations could be attributable to sources other than human-to-human transmission within the geographical setting (i.e., non-local transmission).

**Conclusions:** This study suggests that overall antimicrobial consumption will be, by far, the most powerful driver of future prevalence and that a large fraction of colonizations could be attributed to non-local transmission.

## Introduction

Over 650,000 infections with antimicrobial-resistant bacteria with more than 33,000 attributable deaths are estimated to occur every year in the European Union (1). The rapid spread of extended-spectrum betalactamase (ESBL-) producing Enterobacteriaceae, and the subsequent increase in carbapenem consumption is of global concern (2–5). Local and external interacting sources of colonization mediate this spread in a population. Local sources of colonization include human-to-human transmission within communities that share geographic proximity. External sources of colonization include traveling to regions with high prevalence of colonization with ESBL-producing Enterobacteriaceae (high-prevalence regions), as well as consumption of contaminated food (6–8). Transmission of ESBL-producing Enterobacteriaceae, in particular *Klebsiella pneumoniae*, is known to be enhanced by hospitalizations and antimicrobial consumption (7, 9, 10). Infections caused by *K. pneumoniae* are the third most prevalent among resistant pathogens (1) and a major clinical challenge.

Theoretical and data-driven studies have characterized the sensitivity of drug resistance to differences in the spatial distribution of antimicrobial drug use (11–13). But to which extent antimicrobial consumption and local and external sources of colonization have contributed to the spread of ESBL-producing *K. pneumoniae*, and the magnitude of their effect on the future prevalence of colonization with this pathogen, is not yet understood. This knowledge is however necessary to accurately project the future burden of colonization with ESBL-producing *K. pneumoniae*, and to design public health interventions capable of effectively tackling this process.

The prevalence of colonization with ESBL-producing *K. pneumoniae* in Switzerland is steadily increasing throughout the country, as evidenced by data from the Swiss Centre for Antibiotic Resistance; (ANRESIS). This prevalence is lower in Switzerland than it is in its neighbouring countries, and similar to that in the Netherlands, Norway and Denmark (14, 15). The future prevalence of colonization with ESBL-producing *K. pneumoniae*, and the potential of public health interventions to reduce it, remain uncertain.

This study aimed to project the future prevalence of colonization with ESBL-producing *K. pneumoniae* under different public health interventions by means of a mathematical model calibrated to a population with stable structure and subject to incoming sources of colonization. We did this by means of a mathematical model that reconstructed the course of the prevalence of ESBL-producing *K. pneumoniae* and antimicrobial consumption within hospitals and in the community as observed over 13 years in a Swiss region with stable population structure and detailed in- and out-patients records.

## Methods

### The ANRESIS database and the modelled population

Antibiotic resistance and antimicrobial consumption data were obtained from the Swiss Centre for Antibiotic Resistance database (ANRESIS), which has been previously described in detail (16). In brief, ANRESIS prospectively collects routine and patient-specific antibiotic resistance data representing around 80% of annual hospitalization days across Switzerland (17). Most laboratories gather data from multiple hospitals, ranging from primary- to tertiary-care institutions and from the community (general practitioners). The model developed for this study used data from the Swiss Canton of Valais. This canton is geographically relatively isolated and has a population of approximately 350,000 inhabitants, which remained stable throughout the observed years (18). All 25 healthcare institutions in the Valais participate in ANRESIS, with five public hospitals contributing most of the data. Data have been collected since 2004 using standardized methodology (19).

### Antimicrobial resistance

We used the outcomes of susceptibility tests to ceftriaxone as surrogate for presence/absence of ESBL-production. We considered all clinical samples of invasive (from a usually sterile site) and non-invasive *K. pneumoniae*, and assumed invasive samples to represent clinically significant infections, and non-invasive samples to represent non-clinically significant infections and/or colonization. Clinically significant infections were assumed to lead to antimicrobial therapy. We included susceptibility data collected between 2004 and 2017. Estimations of prevalence excluded repeated tests with the same ceftriaxone resistance results for the same patient within the same year.

### Antimicrobial consumption

Outpatient data in 2017 include around 50% of all of prescribed antibiotics as 56% (69/123) pharmacies in the region reported their data to ANRESIS, runs assumed that it was also the case for the simulation period. Inpatient data included data from all public hospitals. Inpatient antimicrobial consumption was imputed for the smaller, private institutions on the basis of yearly bed-days and accordingly to the consumption in the public hospitals. Data collection spanned between 2006 and 2015 for inpatients and between 2013 and 2016 for outpatients. Consumption data was expressed in defined daily doses (DDD) for the 5th level of hierarchy according to the WHOCC-ATC classification (20) (for example: J01CA; [Penicillins with extended spectrum](http://www.whocc.no/atc_ddd_index/?code=J01CA&showdescription=no)).

The model considers three types of antimicrobial therapy termed *regular, restricted* and *neutral. Regular* and *restricted* types represent no activity and activity against ESBL-producing pathogens, respectively. The *neutral* type represents antibiotics that neither select for nor have a clinical relevant effect on ESBL-producing pathogens, or the effect on ESBL producing pathogens is disputed, such as penicillin/betalactame inhibitor combinations (21–23). **Supplementary Table S1** shows all categories in each group.

### Mathematical model structure

We developed a mathematical model simulating the spread of colonization with ESBL-producing *K. pneumoniae* under the pressure of antimicrobial consumption in two settings, represented by two interconnected models: the hospitals and the community. **Figure 1** summarizes this model structure. Model parameters were set to reflect interactions between settings, and their specific transmission dynamics. **Table 1** shows all model parameters. The model assumes that a dynamic fraction of the population is hospitalized, and captures admissions in hospitals and discharges back to the community by means of hospitalization and discharge rates. Overall rates of antimicrobial consumption are assumed to indicate a response to likely bacterial infections (24). Rates of admission, discharge and antimicrobial consumption were set to reflect data from the Valais region recorded by ANRESIS.

**Table 1.**
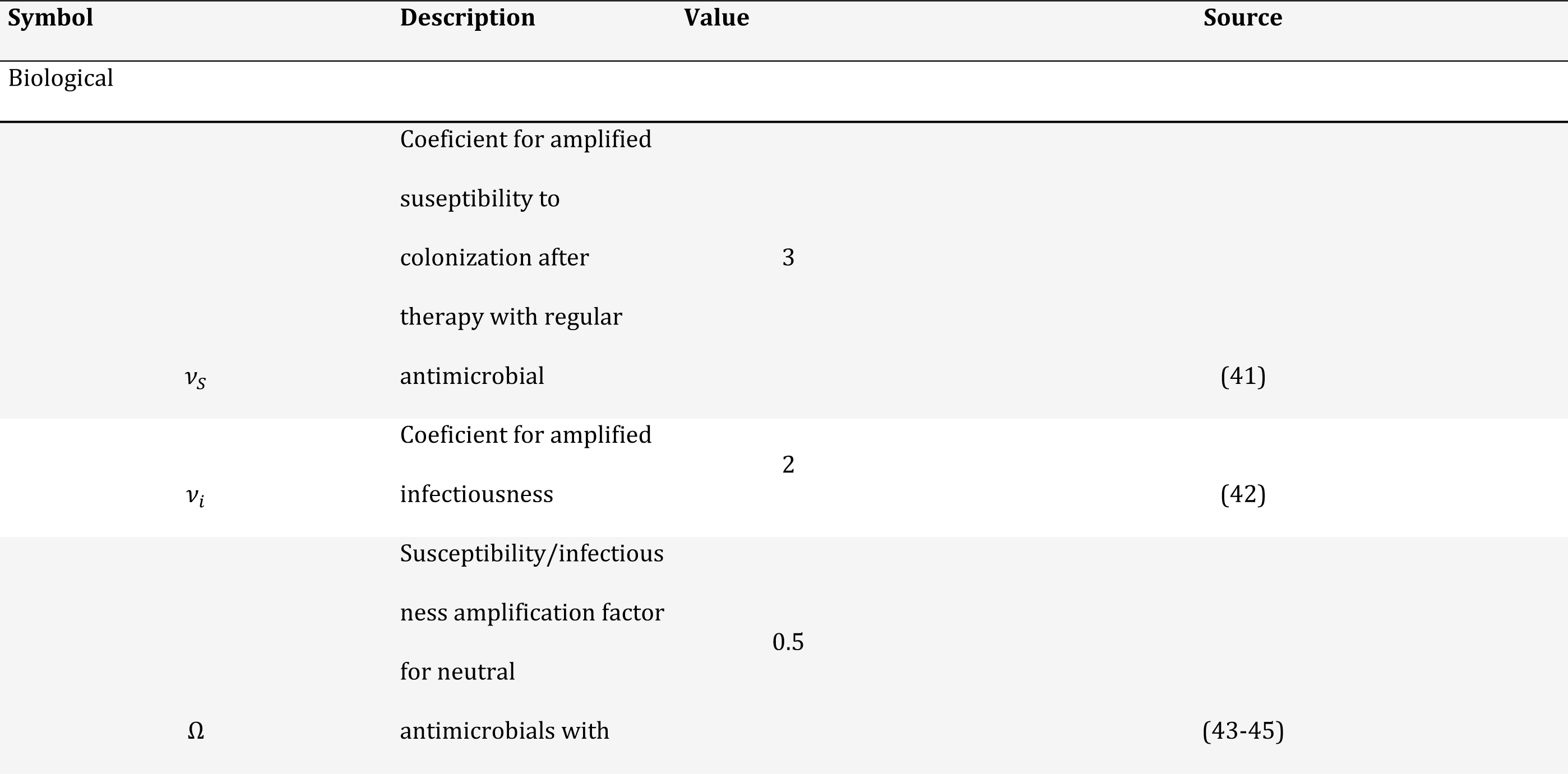
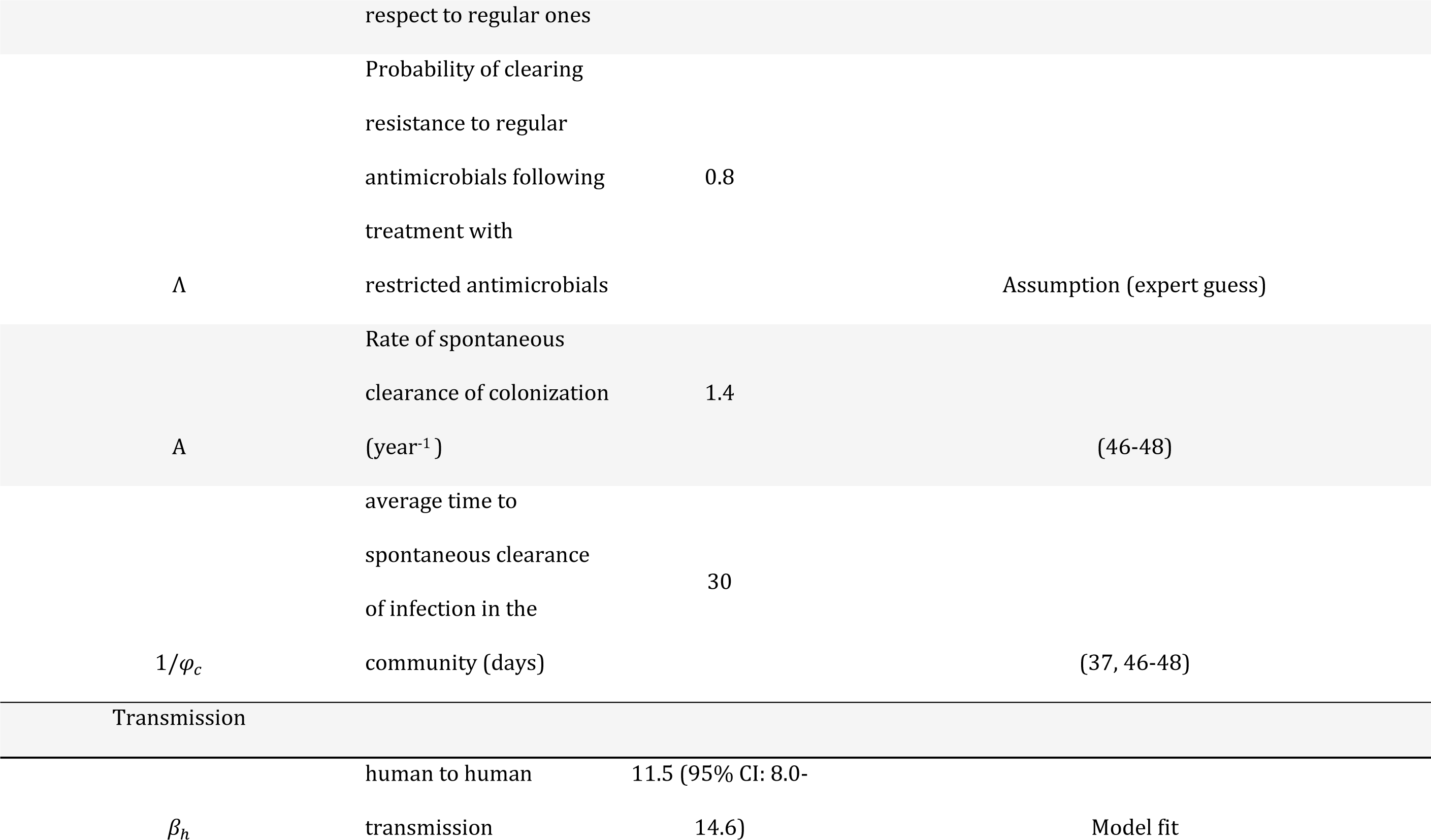
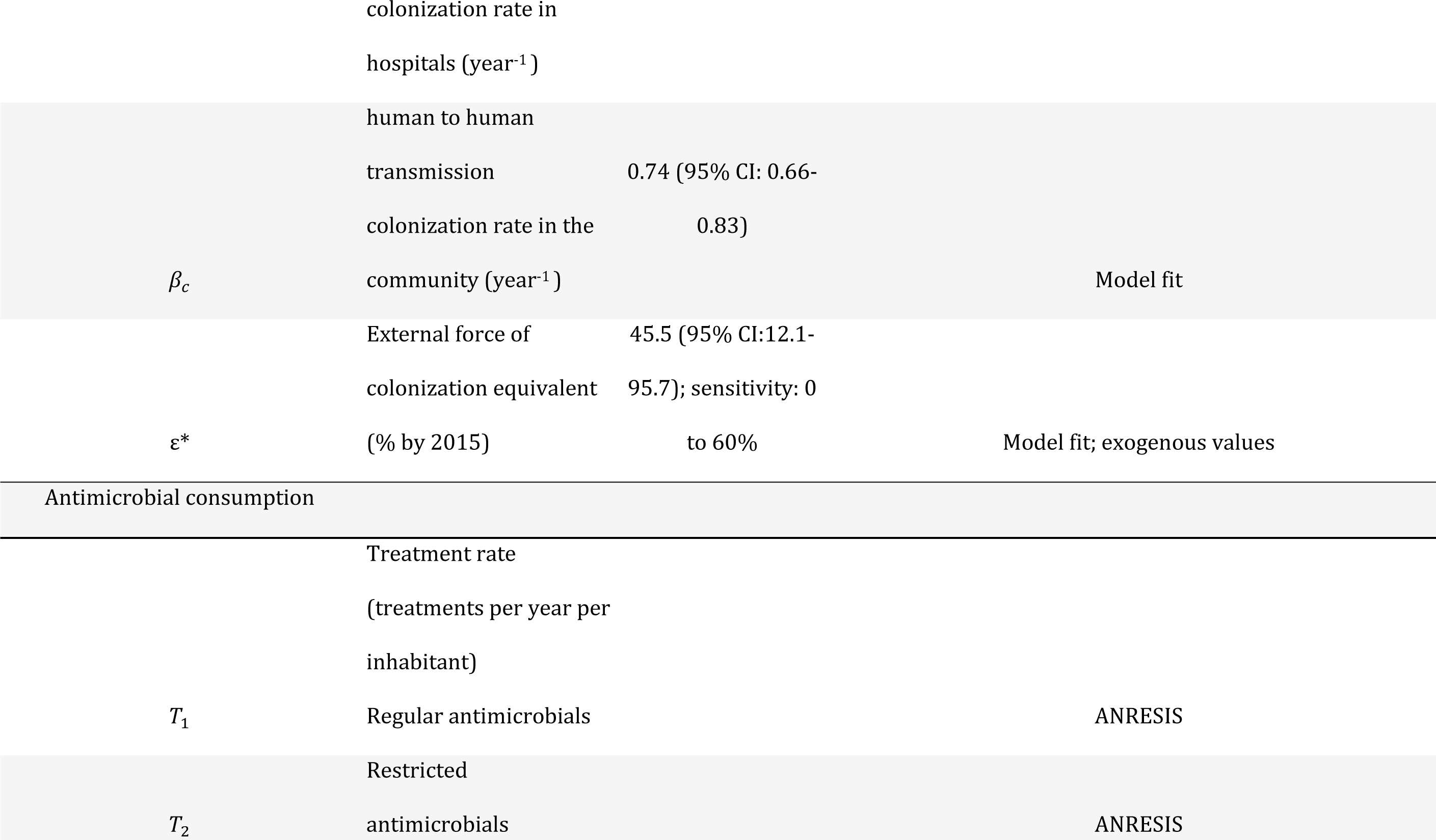
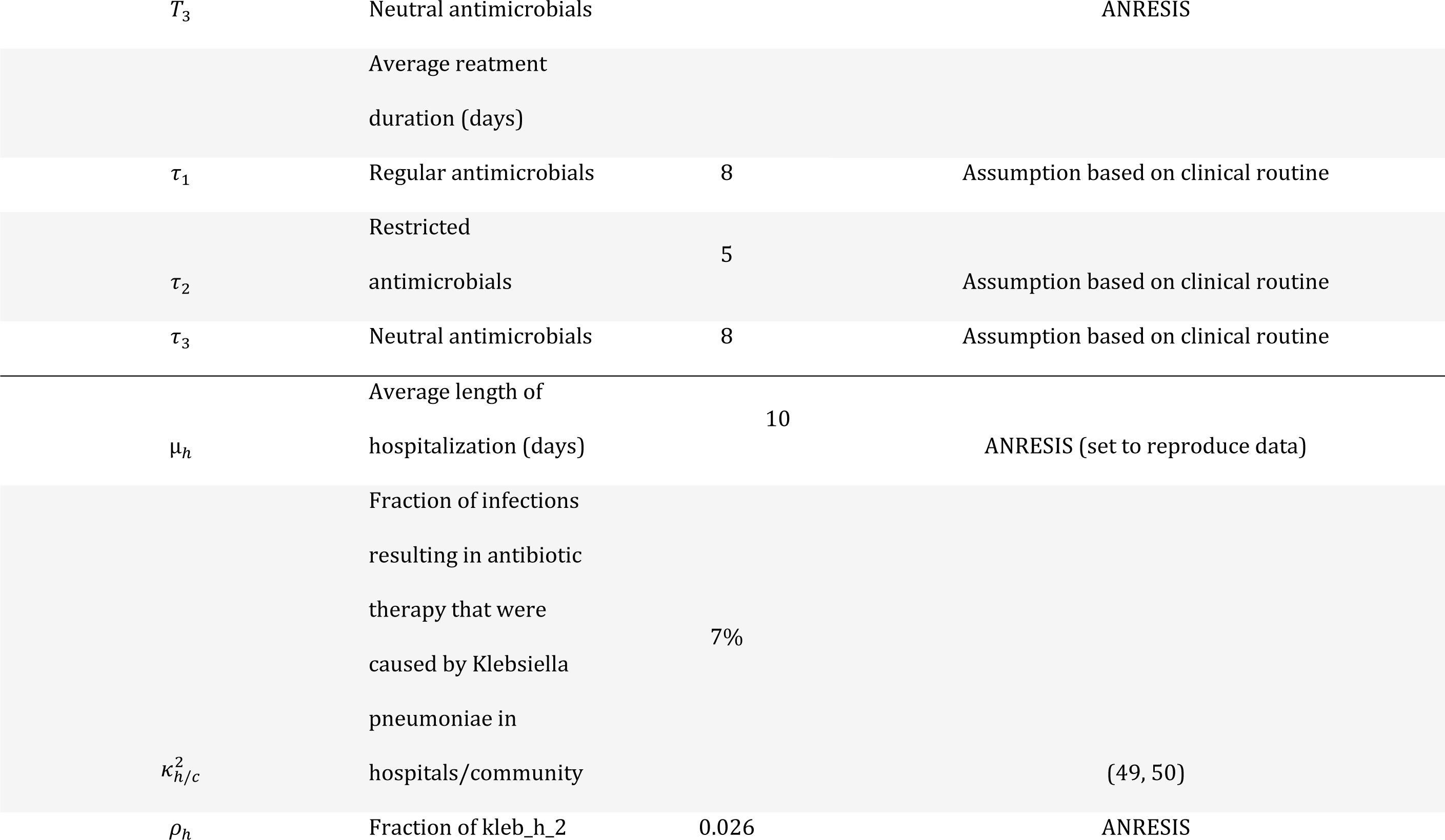
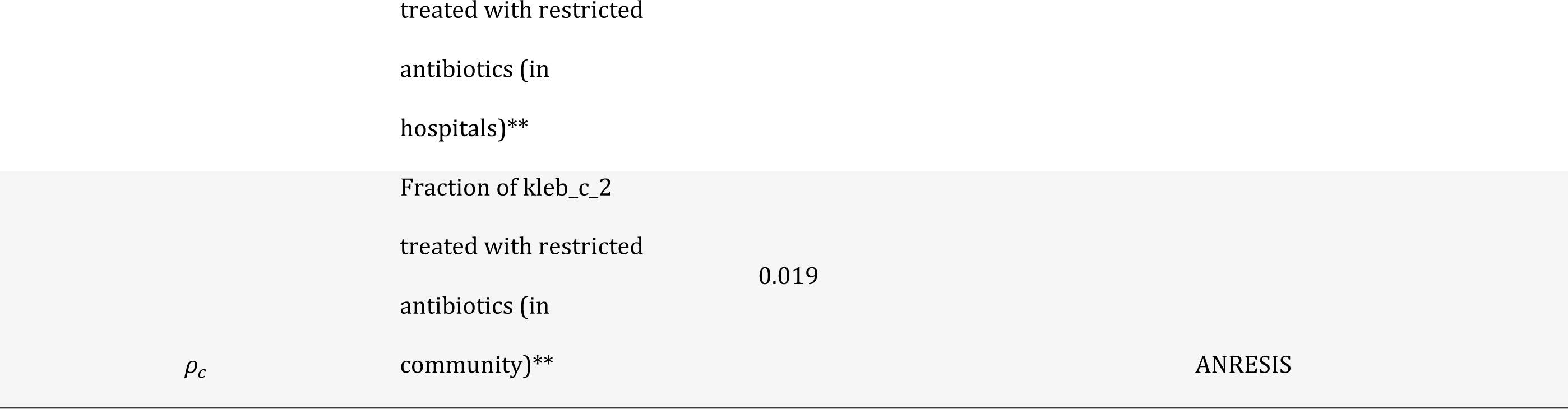
Model parameters definition and sources.

**Figure 1.**
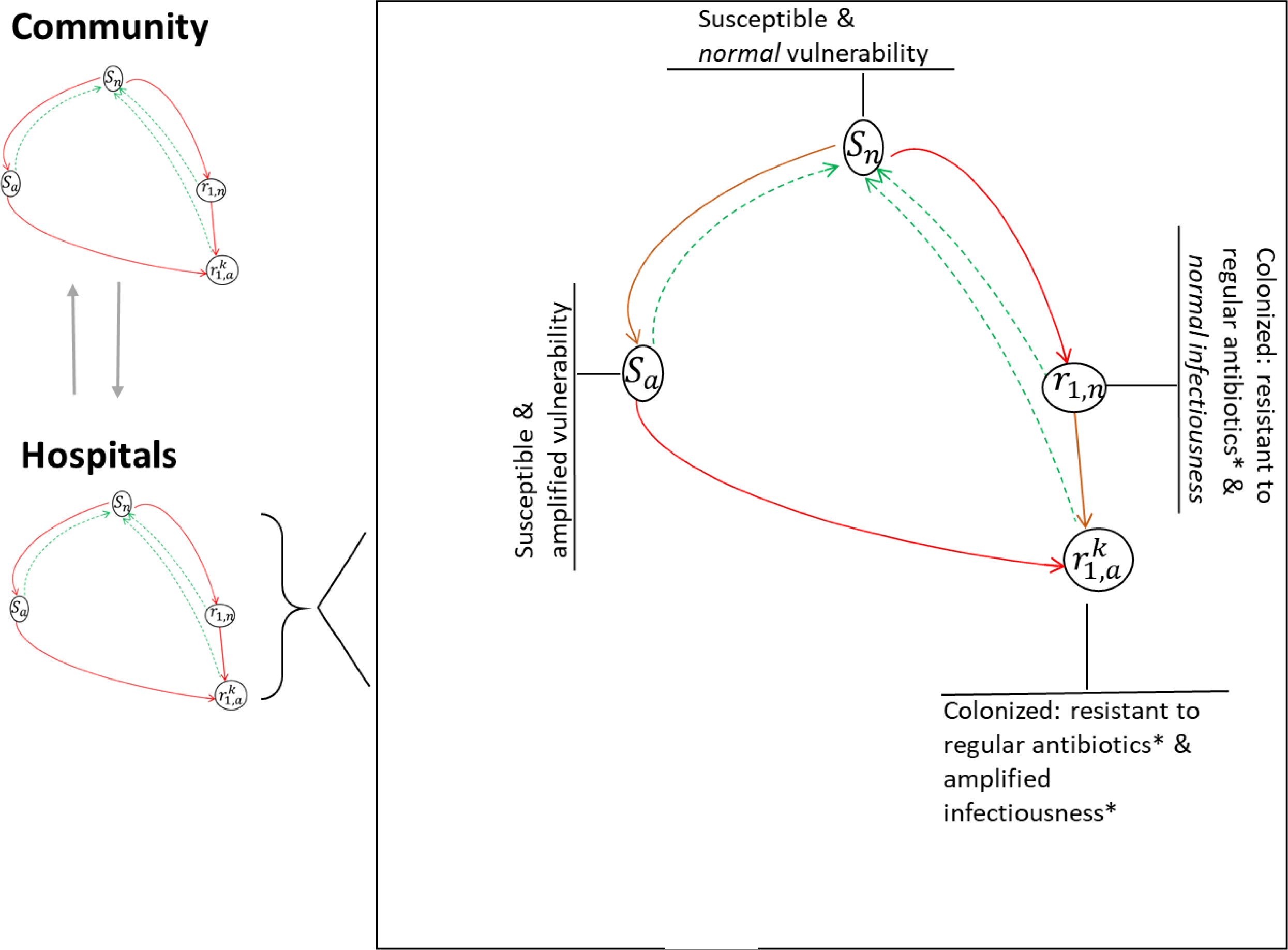
Simplified model structure. The model considers community and hospital settings interconnected by hospitalizations and dismissals. Transmission within each setting is simulated by means of a **core model** with setting specific parameters. In the **core model** (right black box), **uncolonized** individuals can have «normal» (***S***_***n***_) and «amplified» (***S***_*a*_) susceptibility to colonization, reflecting increased risk of colonization associated with antimicrobial therapy (dark, orange arrows). **Colonization** with ESBL-producing *Klebsiella pneumoniae* (red arrows) can occur through human-to-human contact locally (within hospitals and the community) and through external sources (e.g. traveling to high-prevalence areas and contaminated food). **Colonized** individuals are classified into two levels according to their ability to transmit the resistant pathogen: «normal» infectiousness 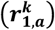 and «amplified» infectiousness (***r***^***k***^) (dark orange arrows). The model explicitly accounts for infections caused by *K. pneumoniae ESBL* with inadequate antimicrobial treatment (*k* = 1, otherwise *k* = 0). *\**ESBL-producing *K. pneumoniae*

### Local transmission

Simulated persons hold a colonization status with ESBL-producing *K. pneumoniae* (colonized vs. non-colonized; compartments labelled *S* and *r*, respectively in **Figure 1**). In the model, the result of local transmission events (e.g. “person-to-person”) is colonization with ESBL-producing *K. pneumoniae*. Transmission can occur in hospitals and in the community i.e., interactions between non-hospitalized persons. The model considers setting-specific transmission rates of the pathogen.

### External force of colonization

This parameter aims to capture colonization through food products (25–27) and from travellers returning from high-prevalence regions (28–30). The model represents these processes through a “force of colonization”, independent of the prevalence within the modelled population. To reflect the increasing number of people travelling to high-prevalence regions, the external force of colonization ε(*t*) was assumed to increase at a constant speed from the year 2000 onwards. To facilitate interpretation of this parameter, we use a more familiar measure, derived from this rate in the results, which we termed “external force of colonization equivalent”. The external force of colonization equivalent was estimated based on the goodness of fit of the mathematical model and labelled ε*. It approximates the proportion of prevalence observed until 2017 attributable to external sources 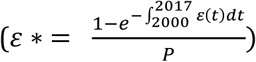 were P is the prevalence measured in 2017 in the community setting).

### Effect of antimicrobial therapy on transmission

The model considered two levels of susceptibility and infectiousness with ESBL-producing *K. pneumoniae*: *normal* and *amplified* (**Figure 1, table 1**). Persons with *amplified* susceptibility (compartment *S*_*a*_ in Figure 1) were more likely to become colonized than those with *normal* susceptibility (compartment *S*_*n*_). An a*mplified* susceptibility status was attained trough antimicrobial treatment. Analogously, persons with *amplified* infectiousness (compartments 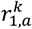) were more likely to transmit ESBL-producing *K. pneumoniae* to others. *Amplified* infectiousness occurred as a result of inadequate antimicrobial treatment. In the model, when a patient colonized with ceftriaxone-resistant *K. pneumoniae* was erroneously treated with regular antimicrobials for an infection caused by this pathogen, such treatment failed and was followed by treatment with restricted antibiotics.

### Model calibration and the role of external force of colonization

We estimated transmission rates for the hospital and community settings by fitting the model to reproduce the data on *K. pneumoniae* ceftriaxone susceptibility in both settings simultaneously. We also iteratively fitted and simulated transmission by assuming external rates of colonization equivalent ε* varying between 0% and 45% in steps of 15%. In an independent analysis, we also estimated the external force of colonization resulting in the best model fit. Model fits were attained by minimizing the sums of squared differences between model outputs and data points weighted to reflect measurement errors in the data.

The model was normalized to a constant population of 100,000 inhabitants.

### Model projections on the impact of public health interventions

We considered public health interventions and changes in clinical routine that would derive in different scenarios of antimicrobial consumption and in hospital transmission. Changes in these variables were modelled as exponential increases and declines, at rates set to reach target levels in 2025.

### Modelled scenarios of antimicrobial consumption

For model projections, we considered hypothetical scenarios of change in: 1) overall consumption of antimicrobials, and 2) consumption of a restricted group of antimicrobials (carbapenems). Antimicrobial consumption was assumed to remain stable at current levels or to reach increases and decreases of 10%, 25% and 50% by 2025.

### Scenarios of in-hospital transmission

We considered hypothetical scenarios where in-hospital transmission remained stable at current levels or reached increases and decreases of 10%, 25% and 50% by 2025.

### Sensitivity analyses on model projections

Because there are no reliable estimates for infectiousness amplification caused by antimicrobial consumption, we assessed the effect on our projections of a range of values for this parameter.

All algorithms, including data processing, statistical analyses, solutions of differential equations, optimizations utilized in model fitting procedures, sensitivity and uncertainty analyses were implemented in R version 3.4.2 (31). In particular, the packages *deSolve* (32), *optim* (33) and *FME* (34) were functional to the outcomes reported in this study.

## Results

Model calibration included data from susceptibility tests performed in 15,137 inpatients and 16,050 outpatients. Observed prevalence of ESBL-producing *K. pneumoniae* varied from 1.4% (95% CI: 0.4 – 3.7%) in 2005 to 10.4% (5.0 – 19.2%) in 2017 in the hospitals setting, and from 2.7% (0.7 – 7%) in 2007 to 2.4% (1.0 – 4.9%) in 2016 in the community setting. Consumption of all types of antimicrobials have increased in hospitals, reaching 55,747, 7,694 and 66,177 defined daily doses (DDD) for the *regular, restricted* and *neutral* types in 2015, respectively. Conversely, in the community, the use of *regular* and *neutral* antimicrobials changed only slightly over time, reaching 245,628 and 521,243 DDD in 2016 respectively, while consumptions of antimicrobials of the restricted type increased by 50% with respect to 2013, reaching 1,150 DDD in the year 2015.

### Model calibration

**Figure 2** shows observed and modelled prevalence of ESBL-producing *K. pneumoniae*, and **Table 1** the values for the fitted parameters. Estimated transmission rate in the hospital setting was 15-fold larger than that in the community setting. Fitted external force of colonization equivalent was 45.5 (95% CI: 12.1-95.7%). Modelled prevalence between 2005 and mid-2017 varied from 0.7% (95% CI: 0.4%-1.5%) to 5.8% (5.3%-9.4%) and from 0.3% (0.1%-0.5%) to 2.8% (2.2%-4.7%) in the hospital and community settings, respectively.

The prevalence data point for 2006 in hospitals (**Figure 2**) corresponds to an atypical jump in prevalence. The circumstances that led to this measurement were unfortunately not documented. We included this point and its corresponding uncertainty without any additional penalty.

**Figure 2.**
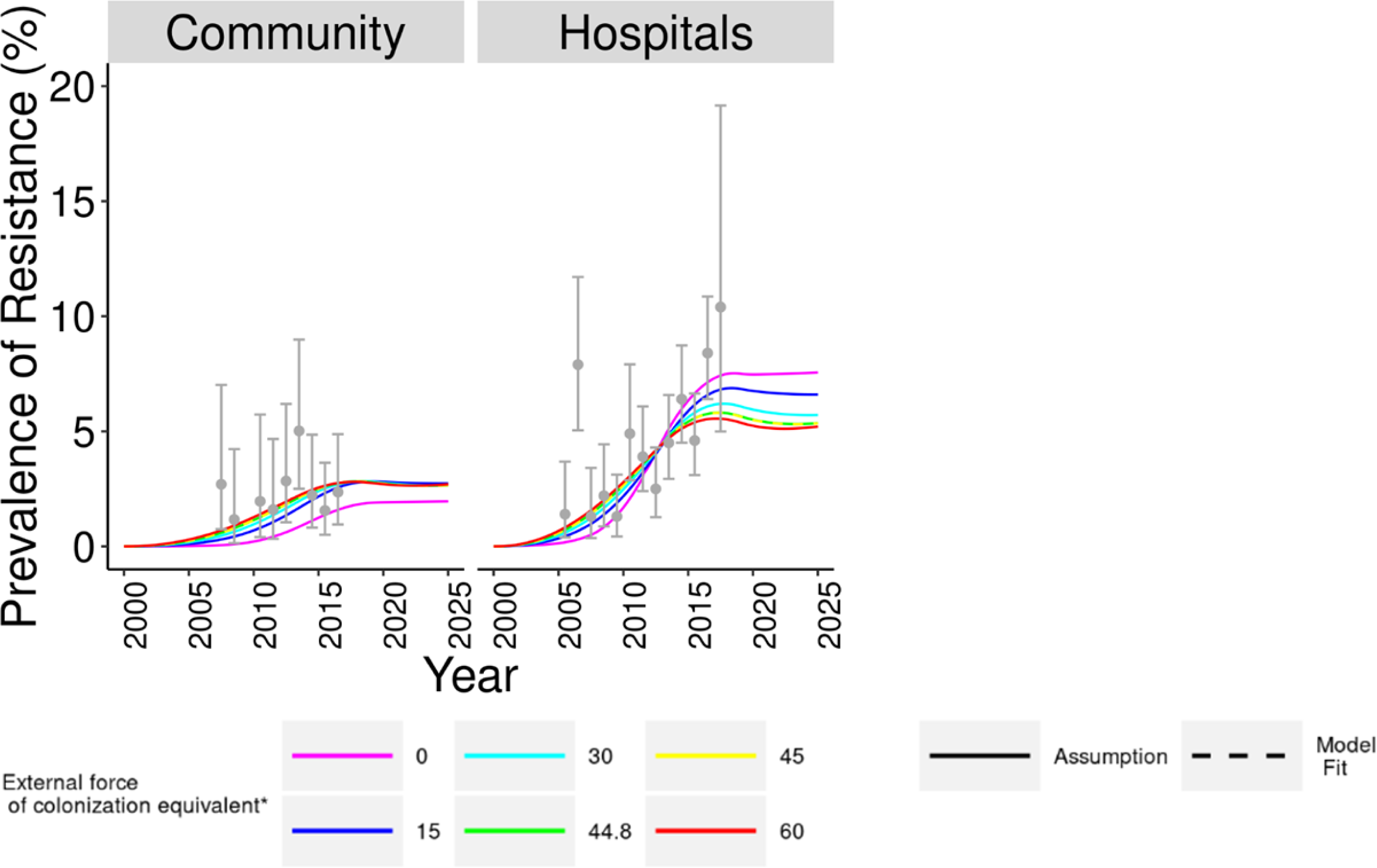
Measured prevalence, model fit and projections of colonization with *ESBL-producing Klebsiella pneumoniae* for a range of external forces of colonization. Data from ANRESIS (grey dots and error bars with 95% confidence intervals) Projected future incidence in hospitals decreased monotonously with increasing external force of colonization. *The external force of colonization equivalent is a proxy for the fraction of observed prevalence by 2017 attributable to external sources.

### Effect of the external force of colonization on prevalence

**Figure 2** also shows the effect of this parameter on model projections. Increasing values for the external force of colonization resulted in lower, stabilizing future prevalence in hospitals. Maximum projected prevalence in hospitals by 2025, obtained by assuming null external force of colonization equivalent, was 7.6% (95% CI: 2.6%-14.1%). Maximum projected prevalence in the community was 2.7% (1.1%-5.4%), with little variation between scenarios of no null external force of colonization. These simulations assumed that antimicrobial consumption and transmission rates remained constant since 2018.

### Model projections on the impact of public health interventions between 2019 and 2025

Simulations assuming changes in overall antimicrobial consumption resulted in the largest changes in projected prevalence. Projected prevalence changed less in simulations considering changes only in carbapenem consumption than in simulations assuming changes in in-hospital transmission rates (**Figure 3**).

**Figure 3.**
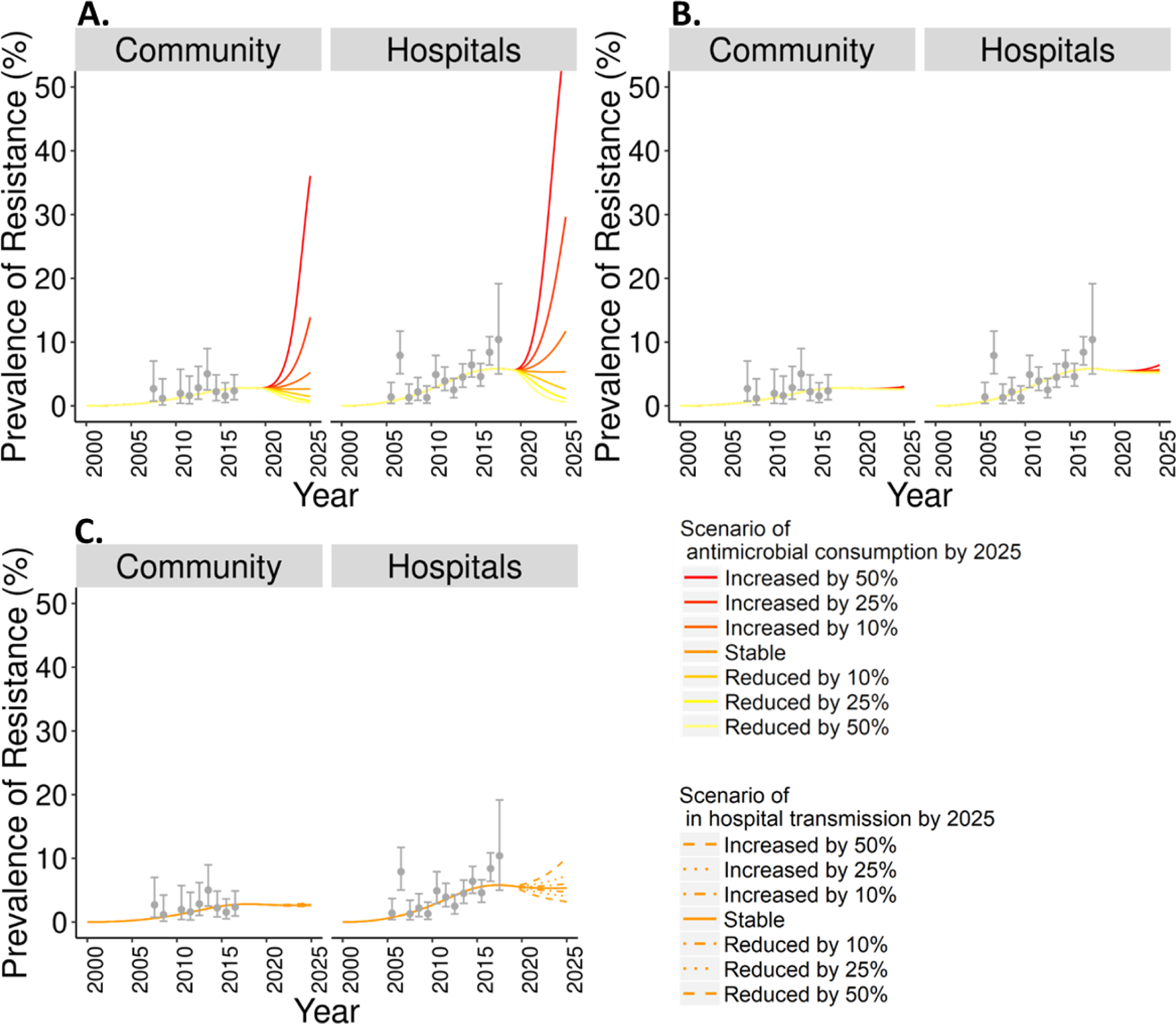
Projections of colonization with *ESBL-producing Klebsiella pneumoniae* for representative scenarios/strategies. Scenarios included changing: antimicrobial consumption (panels A and B), and in-hospital transmission rate (panel C). In Panel A scenarios of antimicrobial consumption included changes in all types of antimicrobials, while in panel B they included only antibiotics of the *carbapenem* class.

### Scenarios of antimicrobial consumption

#### Changes in overall antimicrobial consumption (Figure 3A)

Stable antimicrobial consumption led to almost unchanged prevalence between 2019 and 2025 in both settings. By contrast, future prevalence varied considerably across other scenarios of antimicrobial consumption, with more antimicrobials leading to rapidly increasing future prevalence. A 50% increase in antimicrobial consumption was projected to lead to prevalence of 58.3% (95% CI: 53.6%-64.7%) (10-fold increase from 5.6% (5.1%-9.5%) in 2019) in hospitals and 36.0% (29.2%-43.2%) in the community (13-fold increase from 2.8% (2.2%-5.0%)) by 2025 (**Figure 3A**). Analogously, although at a much lower speed, reductions in antimicrobial consumption led to declining prevalence. A 50% reduction in antimicrobial consumption led to a prevalence of 0.6% (0.4%-1.1%; 89% reduction) in hospitals and of 0.4% (0.3%-0.8%) (84% reduction) in the community by 2025. **Figure S1** shows simulated changes in prevalence by 2025 with respect to 2019 across scenarios of overall antimicrobial consumption.

### Changes in carbapenem-class antimicrobial consumption (Figure 3B)

These simulations, which only varied consumption of carbapenem-class antimicrobials, resulted in changes in prevalence considerably smaller than those reported above did. When carbapenem consumption was set to increase by 50%, projected prevalence was 6.4% (95% CI: 5.9%-11.0%) (13% increase) in hospitals and 3.0% (2.4%-5.2%) (9% increase) in the community. When carbapenem consumption was set to decline by 50%, prevalence declined by less than 9% in both settings (**Figure S2A**).

### Scenarios of in-hospital transmission (Figure 3C)

Prevalence in 2025 almost doubled (5.6%; 95% CI: 5.1%-9.5%) to 10.2% (8.1%-18.4%) in the hospital setting and increased by 9% in the community when we assumed a 50% increase in in-hospital transmission rate by 2025. When in-hospital transmission rate was set to decline by 50%, prevalence dropped by 43% in hospitals and by 13% in the community (**Figure S2B**).

### Sensitivity analyses on the infectiousness amplification mediated by antimicrobials

The results of these sensitivity analyses were similar to those of the main analysis. **Figures S3-6** are analogous to **Figures 2, 3** and **S2** when assuming no amplification (*v*_*i*_ = 1 versus *v*_*i*_ = 2 in the main analysis) and higher amplification (*v*_*i*_ = 3) of infectiousness after antimicrobial treatment with a *regular* agent. Prevalence projected in scenarios without amplification of infectiousness was higher and less sensitive to antimicrobial consumption than that projected in scenarios assuming higher amplification of infectiousness.

## Discussion

We developed a mathematical model that reconstructed the observed course of colonization with ESBL-producing *K. pneumoniae* between 2005 and 2017. This model enabled us to assess the potential impact of changes in clinical practice, including those mimicking public health interventions on the future prevalence of ESBL-producing *K. pneumoniae*. The simulations suggested that with stable rates of antimicrobial consumption and in-hospital transmission, prevalence would stabilize in the near future, as it has been suggested by former studies (35). Expected future prevalence depended on the assumed contribution of non-local transmission (i.e., thought external forces of colonization) during the calibration period. Increasing values for the contribution of non-local transmission imply less local transmission, and led the model to project lower future prevalence in hospitals. This result highlights the urgency to slow down transmission at the local level. Simulated future scenarios showed that the most powerful driver of prevalence will be overall antimicrobial consumption, followed by in-hospital transmission rates. The influence of carbapenem consumption on prevalence stood evidently behind these two across simulations.

In line with published data (36), our model fits estimated transmission rates much higher within hospitals than in the community (15-fold). The best fit also suggested that as much as 46% of the prevalence of colonization measured until 2017 could be attributable to sources external to the modelled population, but uncertainty remained too wide to claim a finding. Data from a recent longitudinal study from the Netherlands suggested a 43% prevalence of ESBL-producing *Escherichia coli* and *K. pneumoniae* among people who recently travelled to high-prevalence regions. In the same study, these travellers accounted for 18% of all cases of colonization with these pathogens (37). Another study that screened retail raw vegetables in Amsterdam found ESBL-producing Enterobacteriaceae in 6% of screened samples (38).These findings suggest that in low prevalence settings, an important share of colonizations with ESBL-producing Enterobacteriaceae could not be prevented by means of local public health interventions. Nevertheless, our model projections showed how three different interventions at the local level could influence the future prevalence of colonization with ESBL-producing *K. pneumoniae*. For example, while an increase of only 10% in overall antimicrobial consumption may quickly trigger escalation of colonization, an equivalent reduction may lead to a sharp decrease in its prevalence.

Carbapenem prescriptions are currently intentionally restricted in routine clinical practice (39, 40). In contrast, our model suggests that further reductions in consumption of the carbapenem-class alone would have a modest effect on the prevalence of ESBL-producing *K. pneumoniae*. Of note, mathematical models of pathogen transmission and at healthcare facility levels have investigated the role of antibiotic restriction and sequential treatment with different types of antimicrobials. Their results counterintuitively suggest that antibiotic restriction may promote resistance instead of hindering it (11, 12). The model was originally formulated to also model resistance to carbapenems. However, the setting we modelled did not have reported cases of carbapenem resistance over the study period. Our study could therefore not model the effects of increase carbapenem consumption on carbapenem resistance. This could limit the potential for generalization of our results to settings with prevalent carbapenemase resistance.

In line with former studies, the model was more sensitive to transmission rates than it was to single antibiotic class restrictions (carbapenem class in our study) (13). Simulations projected that eventual increases in in-hospital transmission rates could lead to considerable rises in prevalence in this setting. Reduced in-hospital transmission was effective at reducing prevalence within hospitals, but its effect on the community setting was relatively modest.

Although our findings remained qualitatively unchanged when confronted with different assumptions regarding increases in infectiousness of a resistant bacteria associated with antimicrobial therapy, the generalized lack of estimates for this parameter in the literature may limit the scope of the values we projected for future prevalence. The sensitivity analyses showed that not only did this parameter, which reflects antimicrobial selection pressure, largely determined the magnitude of the impact of antimicrobial consumption on prevalence; it was also decisive for the values of future prevalence, even in the absence of changes in antimicrobial consumption. Our study therefore suggests that precise estimates of future prevalence would require risk factors research aimed at estimating this parameter.

We used the outcomes of susceptibility tests to ceftriaxone as a surrogate for presence/absence of ESBL-production. However, these tests are imperfect surrogate markers for the occurrence of ESBL resistances, as other mechanism may also contribute to non-susceptibility.

Upon re-parameterization, adapting this model to other resistant pathogens with analogous transmission routes, such as ESBL-producing *Escherichia coli*, should be straightforward.

The model was set to reproduce internally consistent time trends on outcomes of resistance tests, antimicrobial consumption, hospitalization and discharge rates, systematically collected. The implications of our findings regarding the comparative ability of different public health interventions to fight the spread of ESBL-producing *K. pneumoniae* are therefore likely to hold true for other regions, even those with higher resistance levels.

### Implications of findings

These results can help inform public policy on strategies to mitigate the spread of resistant bacteria. Expectations regarding the impact of local interventions may need adjustment to account for constrains derived from potentially high contributions of non-local transmission.

This study suggests that interventions including local or national antimicrobial stewardship programs might be most effective if they aim at reducing overall antimicrobial consumption. Further restricting antimicrobials of the carbapenem class is unlikely to noticeably decrease future prevalence of ESBL-producing *K. pneumoniae*. Therefore, in regions with very low prevalence of carbapenem-resistant pathogens, additional restrictions of carbapenem must be carefully weighed against potential detrimental effects of an initially inappropriate antimicrobial therapy.

## Conclusion

The outcomes of our simulations suggest that public health interventions reducing overall antimicrobial consumption would be considerably more powerful at reducing the prevalence of ESBL-producing *K. pneumoniae* than those reducing in-hospital transmissions, or further restricting carbapenem class antimicrobial consumption.

## Data Availability

Available upon request: http://www.anresis.ch/

## Acknowledgments

We thank all patients and health-care workers for their contribution to data collection within ANRESIS. We thank the public Health Service of the Canton of Valais for their permission to use their data on antimicrobial resistance and consumption for this study. We would also like to acknowledge the members of ANRESIS, the Swiss Centre for Antibiotic resistance, and Swissnoso, the Swiss Centre for Infection Prevention. ANRESIS members are: A. Burnens, Synlab Suisse, Switzerland; A. Cherkaoui, Bacteriology Laboratory, Geneva University Hospitals, Switzerland; C. Corradi, Federal Office of Public Health, Bern, Switzerland; O. Dubuis, Viollier AG, Basel, Switzerland; A. Egli, Clinical Microbiology, University Hospital Basel, Switzerland; V. Gaia, Department of Microbiology, Bellinzona, Switzerland; D. Koch, Federal Office of Public Health, Bern, Switzerland; A. Kronenberg, Institute for Infectious Diseases, University of Bern, Switzerland; S. L. Leib, Institute for Infectious Diseases, University of Bern, Switzerland; P. Nordmann, Molecular and Medical Microbioloy, Department of Medicine, University Fribourg, Switzerland; V. Perreten, Institute of Veterinary Bacteriology, University of Bern, Switzerland; J.C. Piffaretti,

Interlifescience, Massagno, Switzerland; G. Prod’hom, Institute of Microbiology, Centre Hospitalier Universitaire Vaudois, Lausanne, Switzerland; J. Schrenzel, Bacteriology Laboratory, Geneva University Hospitals, Geneva, Switzerland; A. F. Widmer, Division of Infectious Diseases and Hospital Epidemiology, University of Basel, Switzerland;

G. Zanetti, Service of Hospital Preventive Medicine, Centre

Hospitalier Universitaire Vaudois, Lausanne, Switzerland;

R. Zbinden, Institute of Medical Microbiology, University of Zürich, Switzerland.

## Authors contributions

Concept and design of the study: RS, LSV; Data collection: NT, AK, CP; Data analysis: LSV, RS, AA; Model formulation: LSV, RS; Model implementation and analyses: LSV. Drafting of the manuscript: LSV, RS; Interpretation of data and model outcomes, revision of the draft and final approval: All authors

## Conflicts of interest disclosure

### Funding

The study was supported by an institutional grant (CTU Forschungsgrant der Inselgruppe)

## Notes

### Competing Interest Statement

The authors have declared no competing interest.

